# Detection and Genomic Characterization of Heartland and Bourbon Viruses in *Amblyomma americanum* ticks from Nebraska

**DOI:** 10.64898/2026.08.06.26359924

**Authors:** Zach Pella, Joanna Moody, Samantha A. Rodriguez, Sarah Chandler, Halie Smith, Amanda M. Bartling, Kaylee S. Herzog, Sarah Uhm, Sydney R. Stein, Peter C. Iwen, Emily L. McCutchen, Joan L. Kenney, Jeff Hamik, Brent Newman, Joseph R. Fauver

## Abstract

Heartland virus (HRTV) and Bourbon virus (BRBV) are emerging tick-borne arboviruses transmitted by the lone star tick (*Amblyomma americanum*) that have caused dozens of cases of human disease in the United States, including multiple fatalities. Despite their significance, entomological, clinical, and molecular surveillance remains sparse, limiting our understanding of HRTV and BRBV distribution and risk. The Nebraska Department of Health and Human Services and the Nebraska Public Health Laboratory expanded tick-borne pathogen surveillance to include HRTV and BRBV in *A. americanum* ticks beginning in 2024. Here, we report the first detections of HRTV and BRBV in Nebraska and present a multi-segment phylogenetic analysis of complete virus genomes. Using a newly developed amplicon-based whole genome sequencing strategy, we generated complete HRTV genomes from three PCR-positive *A. americanum* pools collected in two counties in eastern Nebraska. Additionally, we generated a complete BRBV genome from a single PCR-positive *A. americanum* pool. A time-calibrated phylogenetic analysis of the L segment containing all publicly available HRTV sequences determined that the 3 genomes from Nebraska form a monophyletic cluster that initially diverged from viruses isolated from Missouri in the early 2000s, corresponding with the expansion of *A. americanum* into Nebraska. A phylogenetic analysis of BRBV segment 2 indicates that the genome from Nebraska sits on a long branch and likely diverged from other genomes sequenced in the early 2010s. Topological concordance across each segment suggests minimal occurrences of reassortment among the HRTV and BRBV genome sequences. These findings document the expansion of HRTV and BRBV to the western margin of the *A. americanum* range and demonstrate the utility of enhanced surveillance and whole genome sequencing for characterizing the spread of tick-borne arboviruses.

## Introduction

The incidence of vector-borne diseases in the United States has been steadily increasing over the last twenty years due to range expansion of vector species, land use changes, introduction of novel pathogens, and recognition of emerging pathogens (1,2). The range of the lone star tick, *Amblyomma americanum*, has expanded to include much of the eastern United States (**Figure 1A**) directly leading to increased risk of transmission for associated pathogens, including arthropod-borne viruses (arboviruses) (3,4). Heartland virus (HRTV) and Bourbon virus (BRBV) are two recently identified tick-borne arboviruses first detected in the central United States within the last 15 years (5). HRTV (*Bandavirus heartlandense*) is a Bandavirus first isolated from two cases in northwest Missouri in 2009 and has subsequently been identified in 14 states causing >60 cases of disease and multiple fatalities (6). BRBV (*Thogotovirus bourbonense*) is a Thogotovirus first identified from a human infection in Kansas in 2014 where HRTV infection was initially suspected, and a total of three human cases have subsequently been identified (7,8). Entomological investigations following initial case detections and characterizations implicated *A. americanum* as the primary vector of both viruses (9–11). Subsequent targeted surveillance has identified both HRTV and BRBV sporadically across the range of *A. americanum* in the United States (**Figure 1B**) (5,9–16). However, HRTV and BRBV have yet to be identified in *A. americanum* populations across much of the species’ distribution, including eastern Nebraska, where *A. americanum* has recently become established and which represents the northwestern edge of the range (17). An incomplete picture of where enzootic transmission is occurring for these important arboviruses hinders our understanding of disease ecology and human risk. In this study, we assessed *A. americanum* ticks collected in Nebraska from 2022-2026 for the presence of HRTV and BRBV to evaluate the potential for human exposure.

**Figure 1.**
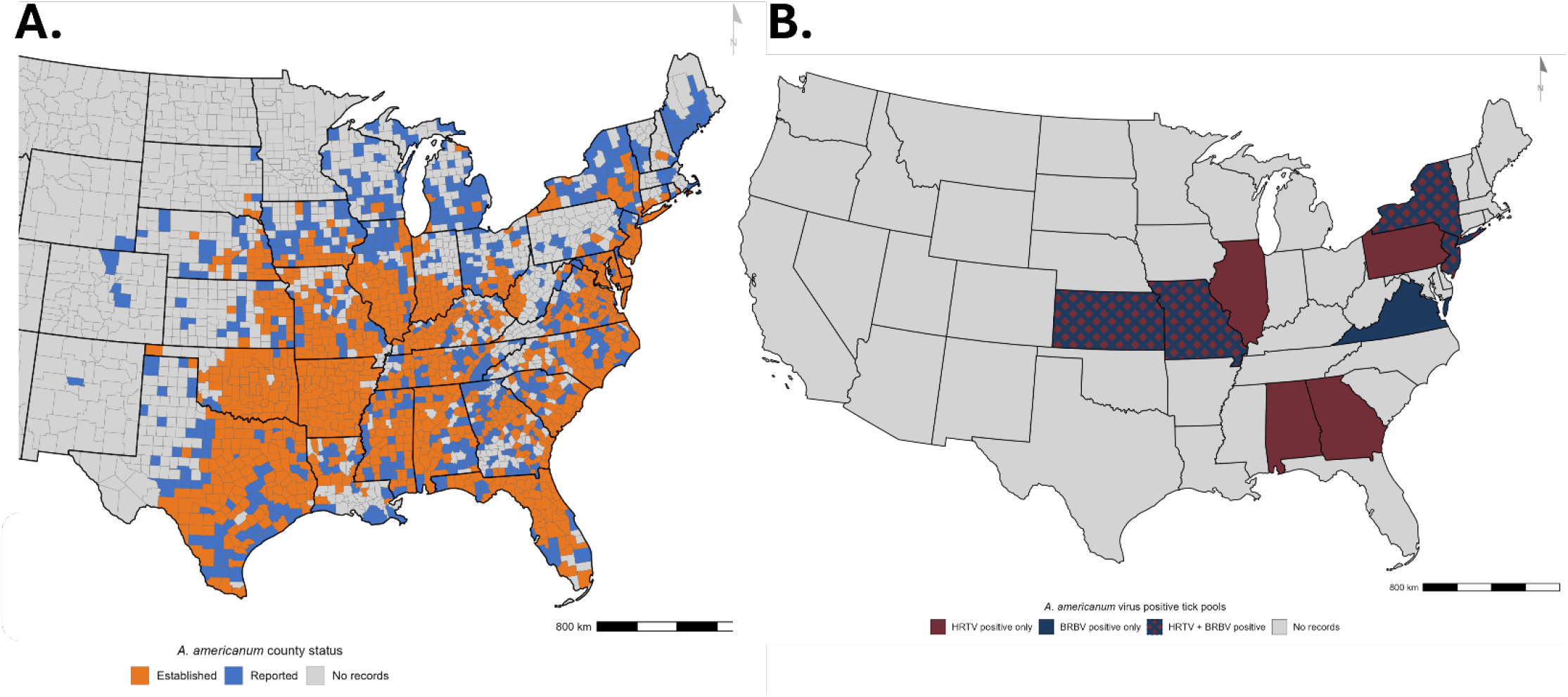
Distribution of *Amblyomma americanum* ticks and HRTV and BRBV detections across the United States. **1A**- Distribution of *Amblyomma americanum* in the United States with county-level occurrence status based on data from the CDC Lone Star Tick Surveillance program (https://www.cdc.gov/ticks/data-research/facts-stats/lone-star-tick-surveillance.html). Counties are categorized as “Established” (orange), “Reported” (blue), or “No records” (light gray). State borders are shown in black; county borders are shown in lighter gray. Map projection: Albers Equal Area Conic (EPSG:5070). **1B**- Geographic distribution of states where Heartland virus (HRTV; burgundy), Bourbon virus (BRBV; navy), or both HRTV and BRBV (burgundy with navy crosshatch) positive *A. americanum* tick pools have been identified, compared to states with no documented HRTV or BRBV positive pools (light gray). Adapted from Dupuis et al. (2023). Map projection: Albers Equal Area Conic (EPSG:5070).

## Methods

### Amplicon Scheme Design and Validation

A PrimalScheme approach was designed for both HRTV and BRBV. PrimalScheme is designed to sequence RNA virus genomes from various sample types containing low viral copy numbers by utilizing a multiplexed overlapping amplicon generation strategy where primer sequences spanning the length of each segment of the virus genome are pooled into two separate PCR reactions (18–20). Sequencing primers for amplicon generation were designed with the online PrimalScheme tool (21). Primers were generated against reference genomes for each segment of HRTV and BRBV with a target melting temperature of 65 °C and amplicon length of 400 basepairs. Accession numbers for each reference genome and primer sequences for each segment can be found in **Supplemental Table 1**. Primers were obtained from Integrated DNA Technologies (IDT) and were reconstituted to 100 μM stocks prior to pooling. For both viruses, primer sequences from each segment were pooled separately at a final concentration of 10μM prior to multiplex amplicon generation reactions.

To validate the PrimalScheme for HRTV and BRBV, we obtained and sequenced viral RNA from five HRTV isolates provided by the Centers for Disease Control and Prevention’s (CDC) Division of Vector-Borne Diseases (DVBD) and RNA from one BRBV isolate provided by BEI Resources (**Supplemental Table 2)**. A total of 8 μL of viral RNA from each isolate was reverse transcribed using LunaScript RT SuperMix (New England Biolabs, NEB) according to the manufacturer’s protocols. Multiplex amplicon generation occurred in two separate reactions (Pool 1 and Pool 2) using the Q5 High-Fidelity Master Mix (NEB) from NEB with the following thermocycler conditions: heat inactivation for 30 seconds at 98 °C, denaturation for 15 seconds at 98°C and annealing for 5 minutes at 65 °C for a total of 35 cycles. Pool 1 and Pool 2 PCR products were combined for each sample and purified with a 1x clean up using KAPA HyperPure Beads (Roche). Purified amplicons were quantified using the Qubit dsDNA HS Assay Kit (Thermo Fisher Scientific) and normalized to 100 ng prior to input into library preparation. Libraries were prepared for sequencing using the Native Barcoding Kit SQK-LSK-109 (Oxford Nanopore Technologies, ONT) according to the manufacturer’s protocols and were sequenced on an ONT MinION R9.4.1 flowcell for 24 hours. Read-level data were basecalled and demultiplexed using the guppy fast basecalling module available through the MinKNOW software. Consensus sequence genomes were generated using the ARTIC Network Field Bioinformatics protocol with a minimum read depth of 20 and maximum total read depth at any position to 200 (20,22). Following consensus sequence generation, the genome of each virus isolate generated using the PrimalScheme approach was compared to the corresponding known genome sequence for each segment independently. Segments were aligned using MAFFT in the Geneious Prime (2025.2.2) software and assessed for segment completeness and single nucleotide polymorphisms (SNPs) (23,24).

### Sample Collection, Processing, and Molecular Detection

Tick samples were collected by the Nebraska Department of Health and Human Services (NE-DHHS) as a part of routine vector-borne disease surveillance (25). Briefly, ticks were collected from select counties across the state using dragging and flagging approaches per CDC guidelines (26). Ticks were transported on dry ice and identified to species and sex using standard morphological keys. *A. americanum* ticks were pooled based on collection date, location, life stage, and sex prior to testing for HRTV and BRBV. Nymphal stage ticks were tested in pools of up to 50 individuals whereas adults were tested in pools of up to 10 individuals. Beginning in the summer of 2024, *A. americanum* ticks were prospectively tested for the presence of HRTV and BRBV at the Nebraska Public Health Laboratory (NPHL).

Pools of ticks were homogenized using a single ball bearing in a tube of Phosephate-Buffered Saline (PBS) and placed on a tissue homogenizer. Homogenate was cleared via centrifugation and nucleic acid was extracted using the MagMAX Viral/Pathogen Nucleic Acid Isolation Kit (Thermofisher) on a KingFisher Flex instrument (ThermoFisher) according to the manufacturer’s instructions. Extracted nucleic acid was tested for the presence of HRTV and BRBV RNA using the CDC RT-qPCR assays targeting the HRTV small segment and BRBV NP gene (segment 5, ORF) (8,9). Retrospective pools of *A. americanum* nymphs and adults collected during 2022-2023 were tested for the presence of HRTV and BRBV at the CDC DVBD. At the DVBD, nymph (≤10 individuals) and adult (≤5 individuals) pools were homogenized using a QIAGEN TissueLyser II with 3 steel ball bearings in 1000 µL of media containing Dulbecco’s Modified Eagle’s Medium (DMEM), sodium bicarbonate, antibiotic-antimycotic solution, and fetal bovine serum. Following homogenization, nucleic acid was extracted as described above. Detection of HRTV and BRBV RNA was performed via RT-qPCR with the same protocol described above on a Bio-Rad CFX-96 system. Positive controls for both viruses were obtained from the CDC Arbovirus Reference Collection (Fort Collins, Colorado). Minimum infection rates (MIR) for HRTV and BRBV were calculated by dividing the number of individual ticks tested for the presence of virus by the number of pools that tested positive, assuming a single individual tick in the positive pool was infected.

### Whole Genome Sequencing and Phylogenetic Analysis

Total RNA from three HRTV positive pools of *A. americanum* nymphs or adults and one BRBV positive pool of adults underwent reverse transcription, amplicon generation, and library preparation as described above using updated reagents, specifically the Native Barcoding Kit SQK-LSK114.24 and FLO-MIN114 flowcells (ONT). Libraries were sequenced for up to 6-hours and raw read data were basecalled using the MinKNOW basecaller guppy specifying the “fast” model. Consensus sequences were generated for each viral segment using the ARTIC Field Bioinformatics protocol described above. Each segment was aligned to the reference sequence genome to assess quality and completeness using Geneious Prime (2025.2.2).

Phylogenetic analysis of HRTV was conducted independently for each of the three genome segments (L, M, and S). Data generated from this study were analyzed in conjunction with corresponding segments from all publicly available HRTV genomic data. Multiple sequence alignments (MSAs) were created using MAFFT and maximum likelihood phylogenetic analysis was conducted using IQ-TREE (27). Interactive phylogenetic visualizations were generated using the Nextstrain platform (28). Custom Nextstrain builds were developed using Snakemake workflows. The pipeline performed the following steps: 1) sequence indexing to create composition indices for filtering; 2) alignment of sequences to reference genomes using MAFFT with gap-filling using N; 3) construction of maximum likelihood phylogenetic trees using IQ-TREE with 14 threads; 4) tree refinement to estimate time-calibrated trees with molecular clock models and outlier filtering at four interquartile ranges from expectation using TreeTime (29); 5) ancestral sequence reconstruction to infer ancestral sequences and mutations; 6) translation of nucleotide sequences to amino acid sequences using GenBank reference annotations; 7) trait inference to reconstruct ancestral geographic states at both country and state levels using an optimized coalescent model with marginal date inference; and 8) export of Auspice-compatible JSON files for interactive visualization. Time-calibrated trees were visualized using Baltic (30). The reference sequences JX005847.1 (L segment), JX005845.1 (M segment), and JX005843.1 (S segment), all isolated from Missouri in 2009 during the original discovery of HRTV, were used to root the trees (6). The pipeline incorporated geographic trait reconstruction for both country and state-level resolution to visualize HRTV diversity across the central and eastern United States. Phylogenetic analysis of BRBV was conducted using the same approach described for HRTV and an independent analysis was conducted for each of the six BRBV genome segments. The earliest available BRBV sequences MH880287 (Segment 1), MH880288 (Segment 2), MH880289 (Segment 3), MH880290 (Segment 4), MH880291 (Segment 5), MH880292 (Segment 6) collected from Missouri in 2013 were used to root the tree.

### Data for Maps

Maps were generated using R statistical software (v. 4.5.0) with the following packages: sf, tigris, ggplot2, and ggspatial. County-level occurrence data for *A. americanum* across the United States were obtained from the CDC Lone Star Tick Surveillance program (https://www.cdc.gov/ticks/data-research/facts-stats/lone-star-tick-surveillance.html). State-level data on HRTV and BRBV positive *A. americanum* tick pools were obtained from Dupuis et al. (2023). Nebraska county-level tick pool collection data were generated from field collections conducted during 2022–2025, and May-June 2026. All maps were projected using Albers Equal Area Conic (EPSG:5070 for national maps; custom Nebraska-centered Albers for the state-level map).

## Data and Code Availability

Code used for consensus sequence generation, phylogenetic analysis, and schemes containing the appropriate reference genomes, .bed files for primer clipping, and folder structure are available at our GitHub page(https://github.com/ZachPella/HRTV-ARTIC-Analysis). All sequencing data and consensus sequence genomes generated in this study are available at the European Nucleotide Archive (ENA) under Project ID PRJEB110868. These data have been cross-listed on NCBI GenBank® and NCBI Sequencing Read Archive and can be found with the same Project ID/Accession number.

## Results

### HRTV and BRBV Primal Scheme Validation

To validate the PrimalScheme approaches for HRTV and BRBV, we generated and sequenced libraries from viral RNA extracted from cell culture isolates and compared them to the publicly available genome sequence. For HRTV, we generated five complete genome sequences from isolates collected in Kansas, Missouri, and Tennessee, including the original isolate from 2009. For BRBV, we generated one complete genome from the original isolate collected in Kansas in 2014 (**Supplemental Table 2**). Genome sequences generated with the HRTV PrimalScheme aligned to the respective genome sequence with 100% nucleotide identity across each segment for all five samples. The average breadth of coverage (i.e., the proportion of the genome sequenced) was 96.5%, with the S segment displaying the lowest average breadth of coverage at 94.8% and the L and M segments at 97.68 and 97.1%, respectively. The genome generated with the BRBV PrimalScheme aligned to the reference genome with 100% nucleotide identity. The average breadth of coverage across six segments was 97.3%, with PB1 segment having the highest coverage at 98.2% and the M protein segment having the lowest at 95.8%.

### Molecular Surveillance of HRTV and BRBV in A. americanum ticks from Nebraska

Analysis of the surveillance data demonstrates a consistent seasonality and distribution of *A. americanum* ticks in Nebraska (**Figure 2A**). The abundance of *A. americanum* across the state was consistently highest in May (37% of total ticks collected) and June (55.8% of total ticks collected) before declining in July. Abundance data for 2026 remain incomplete and are not presented in this study. While *A. americanum* has been detected in western areas of the state, the distribution is largely confined to the Missouri and Platte River corridors in the east (**Figure 2B**). Molecular testing of 898 pools consisting of 7,635 *A. americanum* ticks for HRTV and BRBV revealed low infection rates for both viruses (**Table 1**). A total of three pools tested positive for HRTV by RT-qPCR; a pool of 25 nymphs collected from Richardson County in 2025 (red box, **Figure 2B**) and a pool of 50 nymphs and a pool of 10 male adults collected in Sarpy County in 2026 (blue box, **Figure 2B**). The estimated MIR for HRTV was 0.62/1,000 ticks in 2025, 1.54/1,000 ticks in 2026, and 0.52/1,000 ticks for the entirety of the study. A separate pool of 10 male adult *A. americanum* ticks also collected in Sarpy County in 2026 tested positive for BRBV (blue box, **Figure 2B**). The estimated MIR for BRBV for 2026 is 0.51/1,000 ticks and 0.01/1,000 ticks for the entirety of the study.

**Figure 2.**
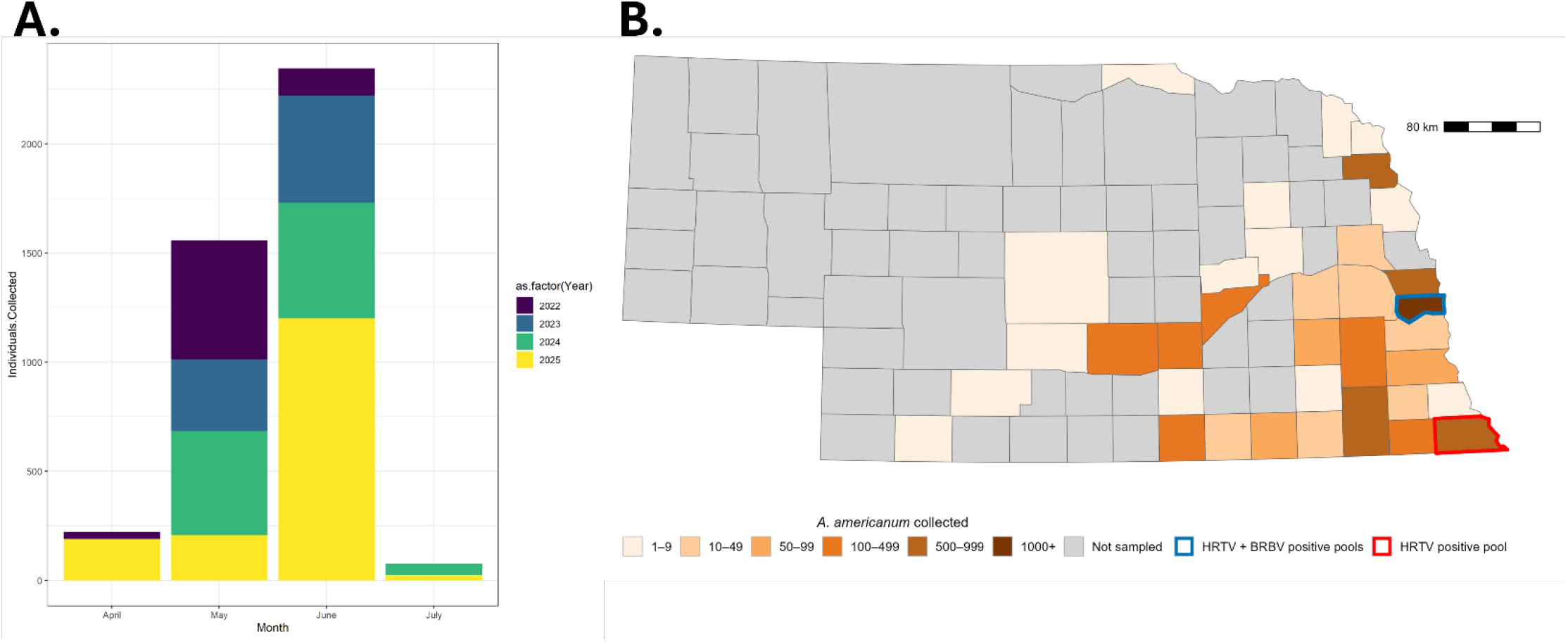
Abundance and Distribution of *Amblyomma americanum* in Nebraska from 2022-2025. **2A**- Abundance and Distribution of *Amblyomma americanum* in Nebraska from 2022-2025. Note that 2026 data is incomplete and not presented. **2A**- Abundance of *A. americanum* in Nebraska presented by year and month of collection. **2B**- Total count of *A. americanum* ticks collected by Nebraska county, 2022–2026, displayed as a sequential orange to brown gradient. Gray counties indicate areas not sampled. Richardson County (outlined in red) marks the location of the HRTV positive *A. americanum* tick pool collected during 2025. Sarpy County (outlined in blue) marks the location where both HRTV and BRBV positive *A. americanum* tick pools were detected during 2026. Projection: Albers Equal Area Conic centered on Nebraska.

**Table 1.** Summary of *A. americanum* survelliance and molecular virological testing data in Nebraska from 2022-2026. ^*^-Data for 2026 remain incomplete

| Year | Pools Tested | Individuals Tested | HRTV Positive Pools | HRTV Minimum Inf Rate | BRBV Positive Pools | BRBV Minimum Inf Rate |
| --- | --- | --- | --- | --- | --- | --- |
| 2022 | 126 | 709 | 0 | 0 | 0 | 0 |
| 2023 | 152 | 817 | 0 | 0 | 0 | 0 |
| 2024 | 217 | 1060 | 0 | 0 | 0 | 0 |
| 2025 | 196 | 1620 | 1 | 0.62 | 0 | 0 |
| 2026* | 108 | 1949 | 3 | 1.54 | 1 | 0.51 |
| **Total** | **799** | **6155** | **4** | **0.650** | **1** | **0.02** |

### HRTV Phylogenetic Analysis

Three high-quality genome sequences were generated from the HRTV-positive pools of ticks detected in this study. HRTV-104 and HRTV-465 had >95% completeness with >50x depth of coverage across all three segments. HRTV-114 had a genome completeness ranging from 75%-95% across all three segments. A time-calibrated phylogenetic analysis for each segment was conducted using publicly available complete genome information to determine the relatedness of the genome from Nebraska to other HRTV samples and to estimate the introduction period. Only the L segment phylogeny is discussed due to observed topological concordance between trees and the phylogeny contains only complete segment sequences. The M and S segment phylogenies can be found in **Supplemental Figure 1**. The metadata for each sample included in the phylogenetic analysis can be found in **Supplemental Table 3**. The analysis of the L segment included 30 publicly available sequences from various US states, spanning 2009 to 2025. Geographic coverage included sequences from Missouri (n=6), Georgia (n=9), New York (n=8), Tennessee (n=1), Kentucky (n=3), Kansas (n=1), Oklahoma (n=1), and Nebraska (n=1). The phylogeny of the L segment reveals clear geographic structure as samples from the same state tended to cluster together (**Figure 3**). The root of the tree was estimated at approximately 1997, with wide 95% confidence intervals on deeper nodes. The most earliest-diverging lineages comprised Missouri sequences JX005847.1 (2009) and OQ688987.1 (2012), consistent with Missouri as the ancestral source population. The HRTV genome sequences from Nebraska form a monophyletic group on a long branch sister to a clade containing samples from Missouri, New York, Tennessee, Kentucky and Georgia. The time to the most recent common ancestor (tMRCA) for the HRTV sample from Nebraska was estimated between 1993-2004.

**Figure 3.**
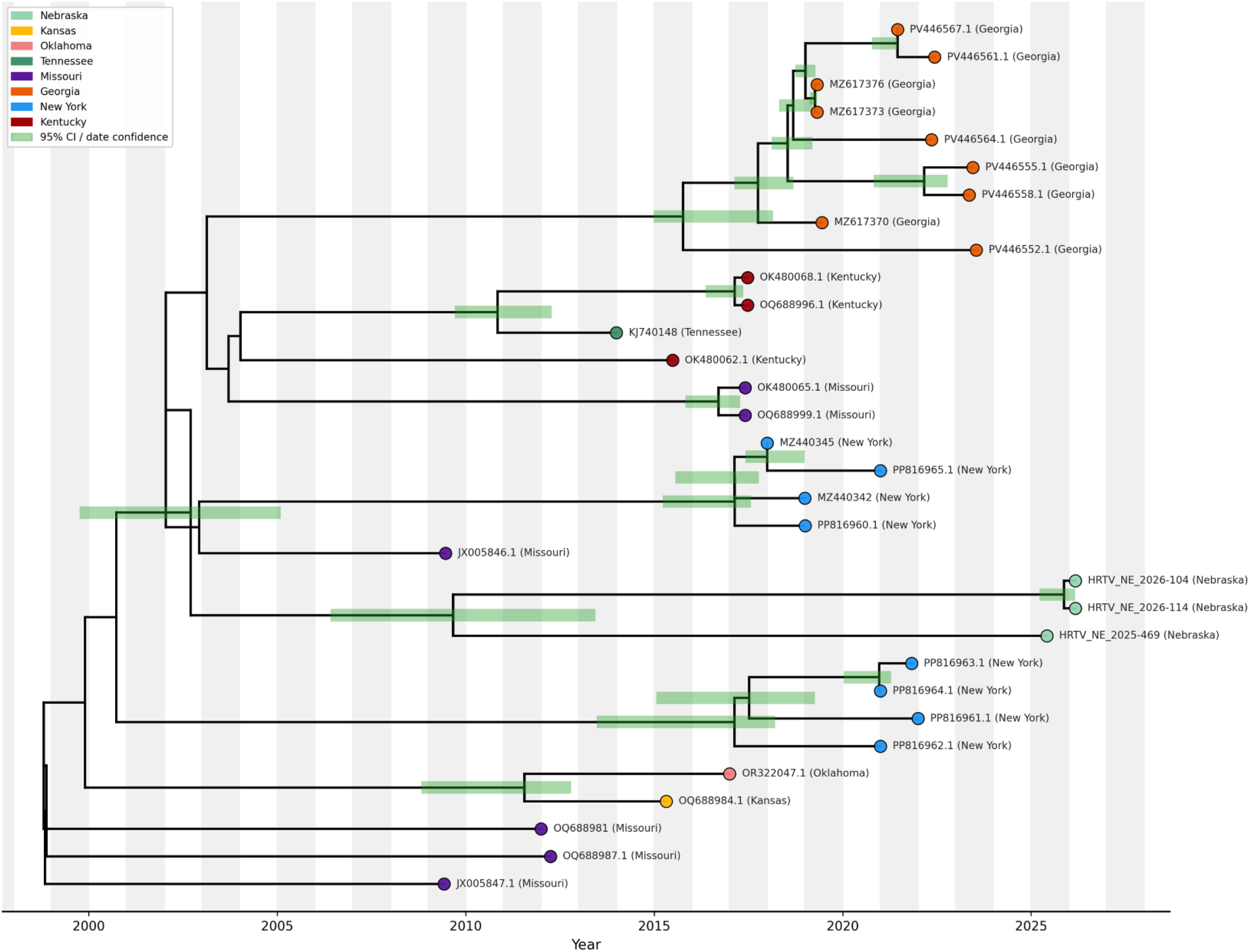
Time-calibrated phylogenetic tree of Heartland virus L segment sequences. Tips are colored by US state of origin. Green bars on internal nodes represent 95% confidence intervals for estimated divergence dates. The Nebraska sequence (UNMC024495, 2025) is highlighted in light blue. The tree was rooted using the reference sequence JX005847.1 (Missouri, 2009) and visualized using Baltic.

### BRBV Phylogenetic Analysis

A single high-quality genome sequence was generated from the pool of BRBV positive ticks, where each of the six segments was >80% complete. A time-calibrated phylogenetic analysis of each segment was conducted using publicly available complete genome information similarly to HRTV. Only Segment 2 (RNA-Dependent RNA Polymerase) phylogeny is disucussed. The phylogeny for the remaining five segments are presented in **Supplemental Figure 2** and metadata for each tree can be found in **Supplemental Table 4**. A total of nine publicly available complete genome sequences of BRBV from the following states were included in the analysis: Missouri (N=3), Kansas (N=2), New York (N=3), and Arkansas (N=1). The sample from Nebraska was recovered on a long branch sister to a clade containing samples from New York and Arkansas (**Figure 4**). Too few genomes are available to reliably estimate tMRCA values.

**Figure 4.**
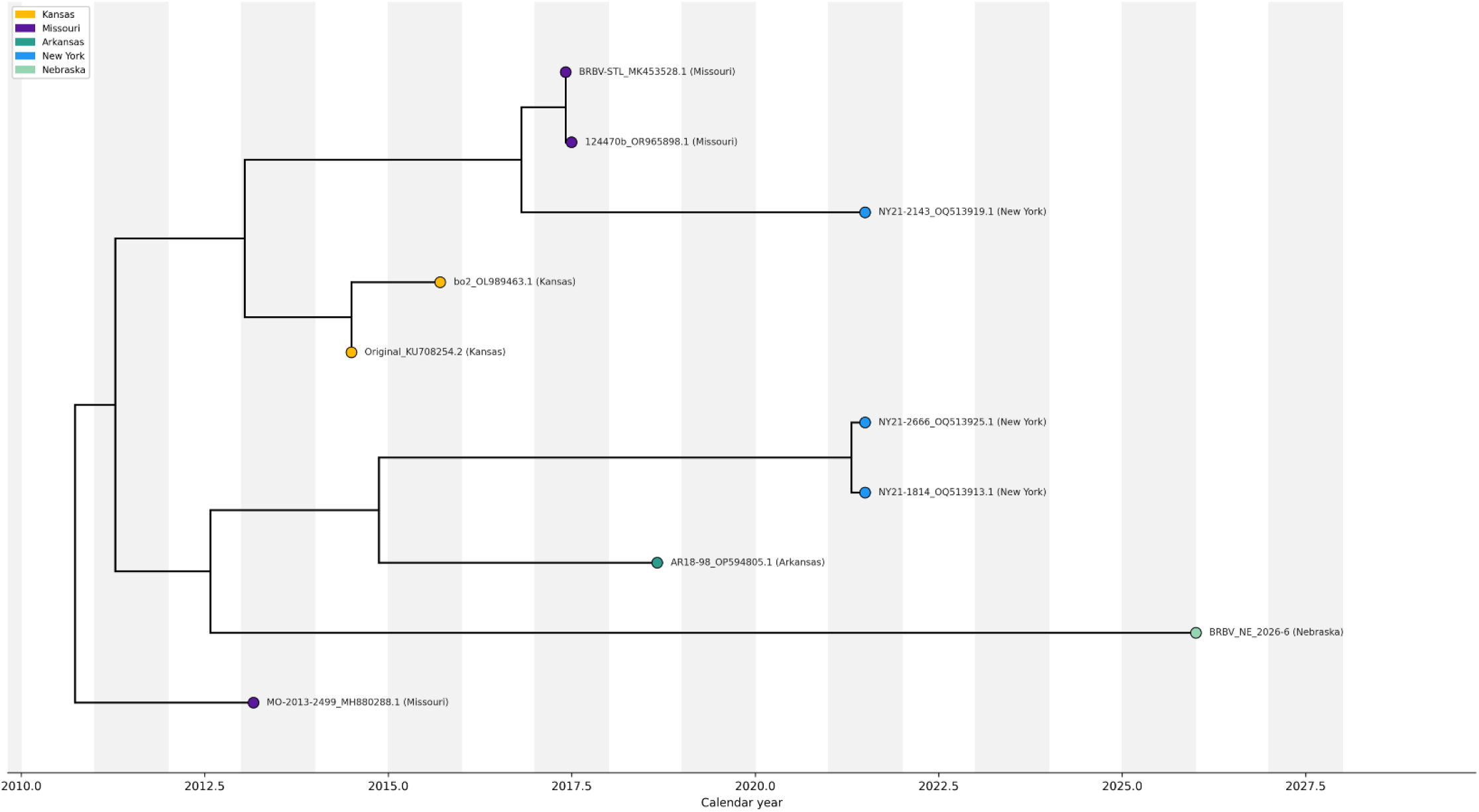
Time-calibrated phylogenetic tree of Heartland virus L segment sequences. Tips are colored by US state of origin. Green bars on internal nodes represent 95% confidence intervals for estimated divergence dates. The Nebraska sequence (UNMC024495, 2025) is highlighted in light blue. The tree was rooted using the reference sequence JX005847.1 (Missouri, 2009) and visualized using Baltic.

## Discussion

Understanding the geographic distribution of ticks and their pathogens is crucial for assessing human risk, particularly where their ranges are expanding. In this study, we provide evidence for HRTV and BRBV detection in *A. americanum* ticks from Nebraska. The majority of the *A. americanum* ticks collected in Nebraska during 2022-2026 were sampled in May and June (92% of all ticks collected) from eastern counties (**Figure 2**) in line with previous studies in Nebraska (17). However, collection effort for routine tick-borne disease surveillance is not uniform across the state. A total of three pools of *A. americanum* ticks tested positive for HRTV, two from Sarpy County in 2026 and one from Richardson County in 2025. The identification in Richardson County is expected given the proximity of the original diagnosed case and description of HRTV from a patient in northwestern Missouri (6). The detection of HRTV in Sarpy County expands the range of HRTV enzootic transmission in Nebraska further north. The only BRBV positive pool identified in the study also came from Sarpy County. The MIR of HRTV in *A. americanum* ticks in this study was 0.52/1,000 ticks (**Table 1**). Similarly low infection rates have been observed in *A. americanum* from Georgia (0.46/1,000 and 0.33/1,000 nymphs) (15,31), Missouri (1.7/1,000) (11), Illinois (9.46/1,000) (13), Alabama (5.7/1,000) (12) and New York (<1.1/1,000) (14). However, much higher infection rates have been observed, including as high as 36.7/1,000 adult male ticks at a field site in St. Louis County, Missouri (16). Infection rates for BRBV were similarly low (0.01/1,000 ticks) in this study and in line with reported infection rates from the few states where BRBV has been detected (32,33).

We developed and validated PrimalScheme assays for both HRTV and BRBV. This amplicon-based approach for whole genome sequencing has the advantage of being target specific, requiring less data to be generated per sample to sequence a complete viral genome, while still being robust to genetic diversity due to the tiled approach and long annealing and extension periods. Using our newly developed PrimalScheme approach for HRTV and BRBV, we generated the first complete HRTV and BRBV genomes from Nebraska. Other PrimalScheme-based assays exist for generating complete HRTV genomes and they were not compared during this study (31). Our time-calibrated phylogenetic analysis of HRTV placed the three genomes from Nebraska in a monophyletic group on a long branch that is most closely related to a sample from Missouri with an estimated tMRCA between 2001-2007 (**Figure 3**). However, the tMRCA estimates from the M and S gene phylogenies are dated further into the 1990s with wider confidence intervals (**Supplemental Figures 1 & 2**).

Given that *A. americanum* was first detected in southeastern Nebraska in 1987 and became established shortly thereafter (17), and the genomes from Nebraska have diverged substantially, our phylogenetic analysis suggest that HRTV was likely introduced to the state as *A. americanum* became established and has maintained an enzootic transmission cycle following introduction. However, the wide confidence intervals of the tMRCA demonstrate a need for increased sampling of HRTV in Nebraska and across the range of *A. americanum* in the United States to better infer phylodynamics and transmission patterns. There are substantially fewer publicly available BRBV genomes for analysis, resulting in insufficient data to reliably estimate a tMRCA for the Nebraska genome (**Figure 4**). Similarly, more genomic data is needed from BRBV to better infer transmission dynamics.

Collectively, the results of this study indicate that HRTV and BRBV are present in *A. americanum* ticks in Nebraska and expand the known distribution of HRTV and BRBV further north and west. While infection rates for both HRTV and BRBV in *A. americanum* in Nebraska remain low, additional studies, including serological surveys of wildlife and humans in the area, need to be conducted prior to estimating risk of transmission to humans. Studies in Georgia and Missouri have identified HRTV consistently at the same sites sampled over multiple years, indicating that HRTV can maintain focal persistence in an enzootic cycle resulting in “hot spots” of human risk (16,31). The cluster of the three HRTV genomes from Nebraska suggests that Nebraska may experience a similar “hot spot” phenomenon, therefore surveillance should continue in eastern Nebraska to determine if HRTV will persist in the area. The presence of HRTV and BRBV in tick populations should inform local public health and healthcare practitioners about potential risk of human infection. This study demonstrates the utility of enhanced entomological surveillance and whole genome sequencing for characterizing the spread of understudied tick-borne arboviruses.

## Acknowledgments

We thank individuals at the CDC DVBD and Dr. Alex Ciota and Dr. Alan Dupis at the Wadsworth Center New York State Health department for providing HRTV RNA for validating the PrimalScheme approach. Ticks used in this study were collected and tested for the presence of HRTV and BRBV as a part of a CDC Epidemiology and Laboratory Capacity grant to NE-DHHS and NPHL. The following reagent was obtained through BEI Resources, NIAID, NIH: Bourbon Virus, Original, NR-50132. This work was partially supported by Cooperative Agreement Number NU60OE000104 (CFDA #93.322), funded by the CDC of the US Department of Health and Human Services (HHS) providing salary support to ZP through the Association of Public Health Laboratoires (APHL). The study is solely the responsibility of the authors and the results do not necessarily represent the official views of APHL, CDC, HHS or the US Government.

