## Supplemental Figures for "Detection and Genomic Characterization of Heartland and Bourbon Viruses in *Amblyomma americanum* ticks from Nebraska"

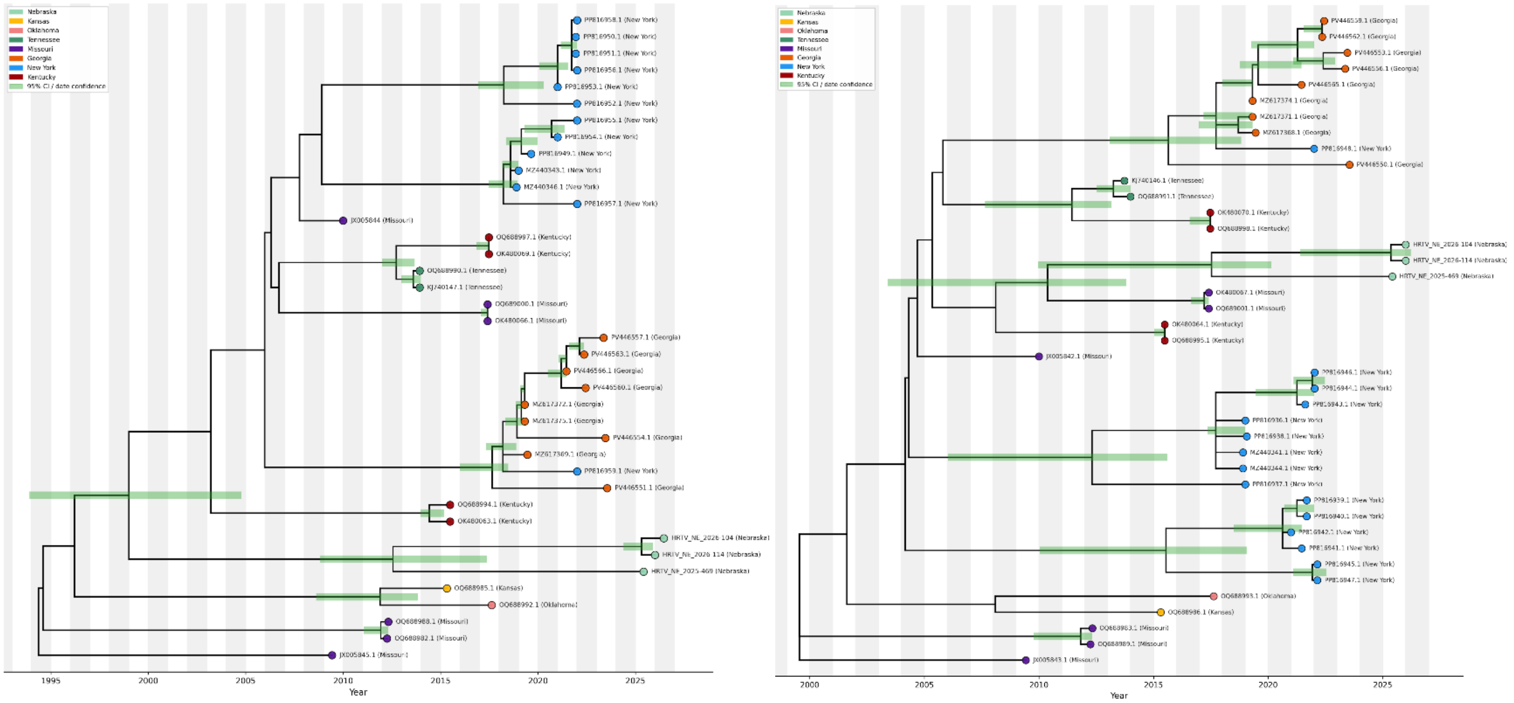


**Supplemental Figure 1. Time-calibrated phylogenetic tree of Heartland virus M and S segment sequences.** Tips are colored by US state of origin. Green bars on internal nodes represent 95% confidence intervals for estimated divergence dates. The Nebraska sequences are highlighted in light green.

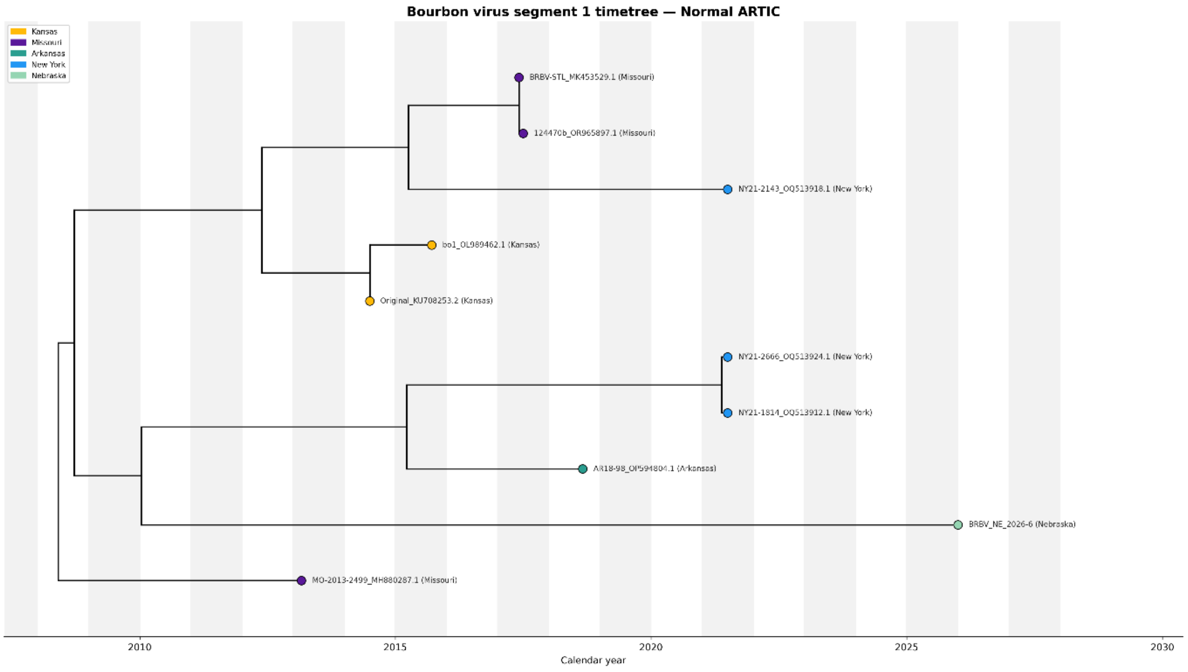

**Bourbon virus segment 3 timetree — Normal ARTIC**

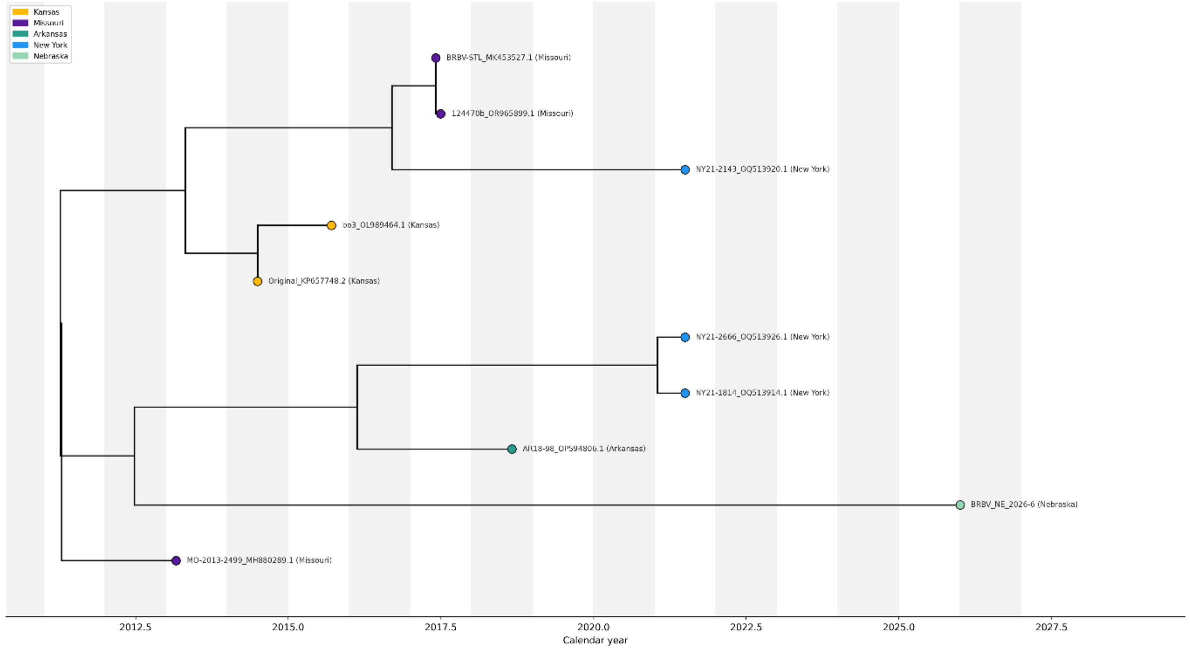


**Bourbon virus segment 4 timetree — Normal ARTIC**

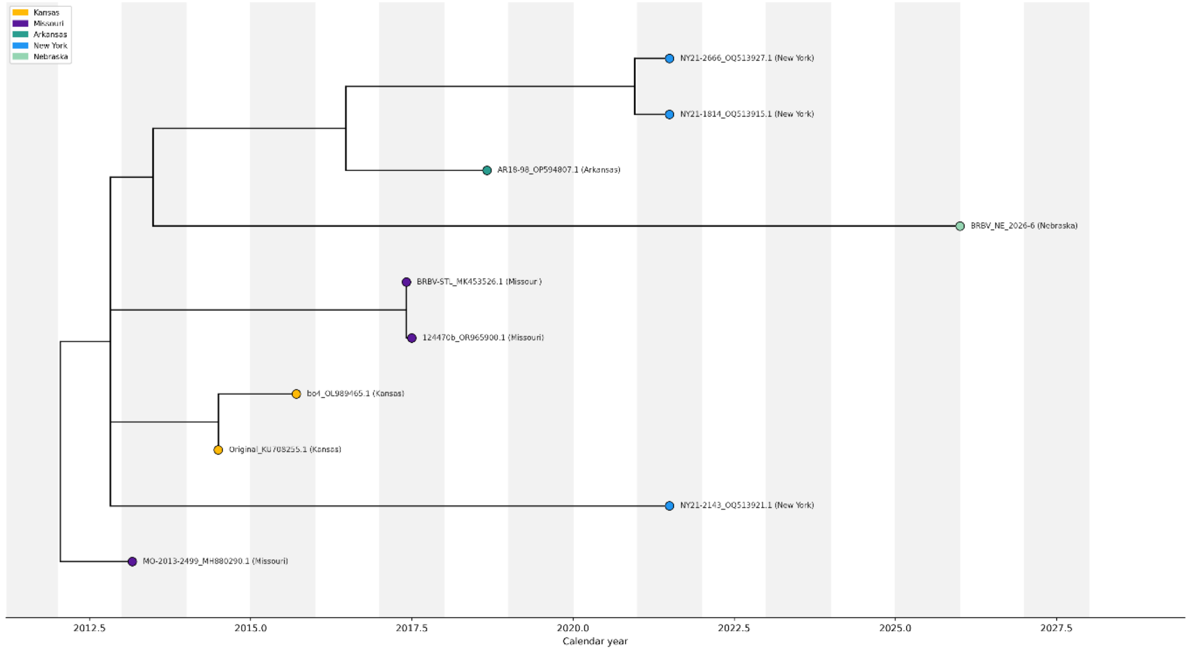

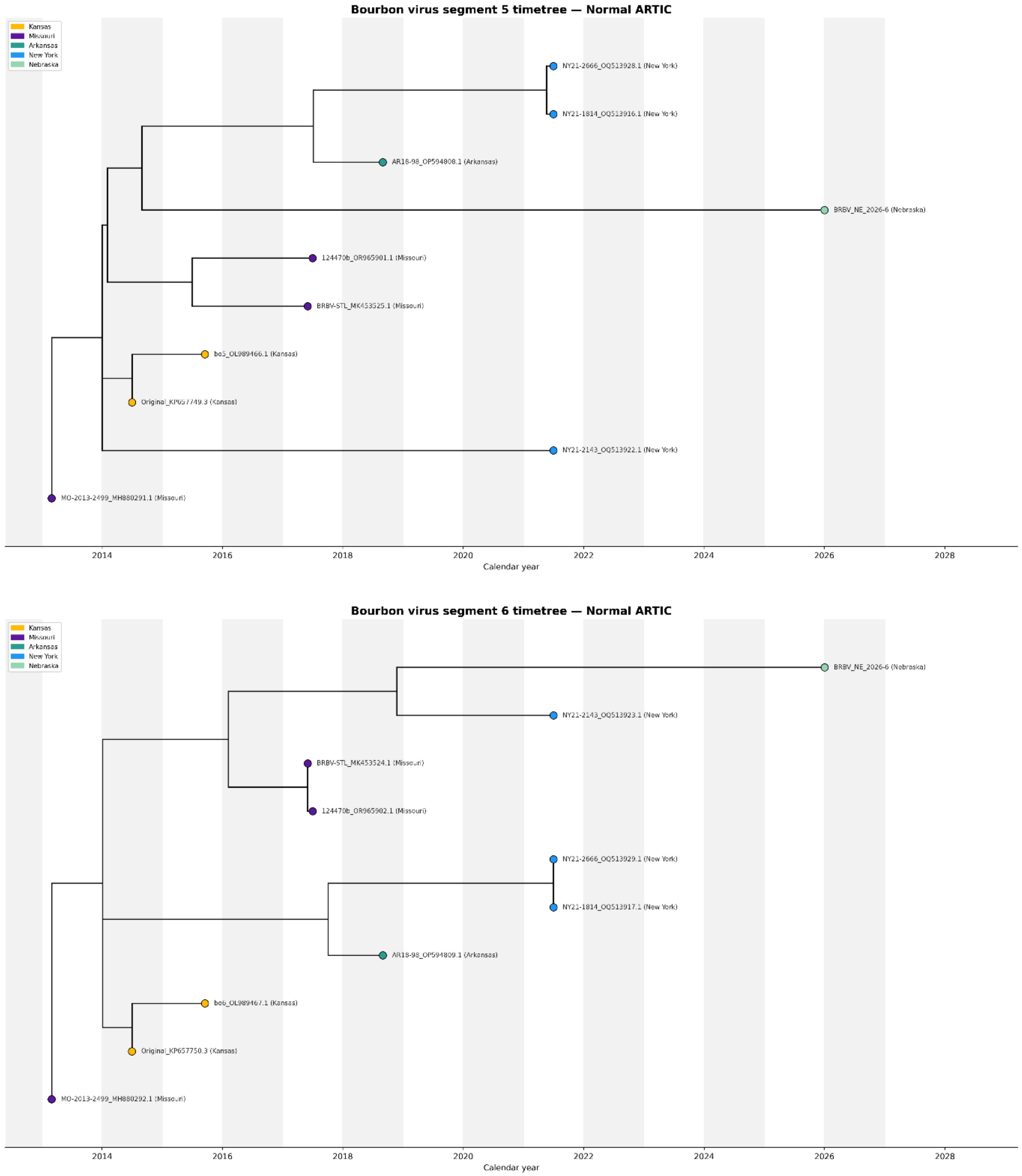


**Supplemental Figure 2. Time-calibrated phylogenetic tree (timetree) of BRBV segments 1,3,4,5, and 6 sequences.** Tips are colored by US state of origin. Green bars on internal nodes represent 95% confidence intervals for estimated divergence dates. The Nebraska sequence (UNMC024495, 2025) is highlighted in light greeb.
